# THEMIS: A Framework for Cost-Benefit Analysis of COVID-19 Non-Pharmaceutical Interventions

**DOI:** 10.1101/2022.04.09.22273656

**Authors:** Dimitris Bertsimas, Michael Lingzhi Li, Saksham Soni

## Abstract

Since December 2019, the world has been ravaged by the COVID-19 pandemic, with over 150 million confirmed cases and 3 million confirmed deaths worldwide. To combat the spread of COVID-19, governments have issued unprecedented non-pharmaceutical interventions (NPIs), ranging from mass gathering restrictions to complete lockdowns. Despite their proven effectiveness in reducing virus transmission, the policies often carry significant economic and humanitarian cost, ranging from unemployment to depression, PTSD, and anxiety. In this paper, we create a data-driven system dynamics framework, THEMIS, that allows us to compare the costs and benefits of a large class of NPIs in any geographical region across different cost dimensions. As a demonstration, we analyzed thousands of alternative policies across 5 countries (United States, Germany, Brazil, Singapore, Spain) and compared with the actual implemented policy.

Our results show that moderate NPIs (such as restrictions on mass gatherings) usually produce the worst results, incurring significant cost while unable to sufficiently slow down the pandemic to prevent the virus from becoming endemic. Short but severe restrictions (complete lockdown for 4-5 weeks) generally produced the best results for developed countries, but only if the speed of reopening is slow enough to prevent a resurgence. Developing countries exhibited very different trade-off profiles from developed countries, and suggests that severe NPIs such as lockdowns might not be as suitable for developing countries in general.

## 1 Introduction

In the last 18 months, the world has been facing one of the biggest health crises in a century – the COVID-19 pandemic. Starting from the initial outbreak in Wuhan (Hui et al. 2020), the disease, caused by the SARS-CoV-2 coronavirus, quickly swept around the globe. As of May 2021, the pandemic took over 3.3 million lives, while new hotspots continue to emerge.

To curtail the spread of SARS-CoV-2 and limit its detrimental humanitarian impact, governments around the world enacted unprecedented non-pharmaceutical interventions (NPIs), ranging from social distancing and mask-wearing to complete lockdowns. Despite their proven effectiveness in reducing transmission, these NPIs incur significant cost on the society. Restrictions on mass gatherings and travel have proven to be detrimental to many industries, greatly reducing economic output while also causing mass unemployment. The travel industry alone lost 3.8 trillion USD in 2020 (WTTC 2021), while the International Labour Organization estimates that in total 114 million full-time jobs were destroyed worldwide (Monitor 2020). Beyond their significant economic cost, more severe measures, such as lockdowns, also incur a large humanitarian cost. The isolation and confinement induced by these policies have caused a sharp rise in depression, anxiety, and PTSD worldwide (Xiong et al. 2020). Researchers have also raised concerns about long-term effects of NPIs such as closing schools, worrying that there will be a decline in the long-term educational attainments.

Therefore, there is significant debate both among the public and the scientific community on whether the benefits of the implemented measures outweigh their cost, and what, if any, better alternatives could have been used. However, quantifying costs from multiple social and economic dimensions is notoriously difficult. To identify the outcome under alternative policies, one also needs to be able to quantify the impact of different NPIs on the spread of COVID-19 and simulate the counterfactual pandemic.

Given these challenges, it is perhaps not surprising that despite global interest, there is relatively little literature that tackles this question holistically. Previous cost-benefit analyses were primarily focused on quantifying the cost-benefit tradeoff for the actual implemented policy (Broughel and Kotrous 2021) or comparing two specific NPIs (e.g. lockdown vs reopening) (Layard et al. 2020, Shlomai et al. 2021) with emphasis on the direct GDP impact of the NPIs (e.g. Rowthorn and Maciejowski 2020, Miles et al. 2021, Gros et al. 2020, Zhao et al. 2021). In contrast, we would look at the pandemic retroactively across the globe. We utilize the realized experience of the COVID-19 pandemic to construct a data-driven framework, THEMIS, that allow us to conduct a cost-benefit analysis of the actual sequence of NPIs implemented in any region across the history of the pandemic, and compare with a wide range of alternatives that could have been implemented. To simulate the effect of the pandemic under different sequence of NPIs (policies), we utilize a recent, policy-driven compartmental epidemiological model called DELPHI (Differential Equations Lead to Predictions of Hospitalizations and Infections). DELPHI extends classical compartmental models to capture key features of the COVID-19 pandemic: (i) under-detection due to limited testing, (ii) governmental and societal response to the pandemic, and (iii) declining mortality rates (Li et al. 2020). Since its inception in April 2020, DELPHI has been applied to more than 200 countries and regions worldwide, producing predictions at the country level and at the state/province level for a few countries. The DELPHI forecasts have been incorporated into the central ensemble at the US Center for Disease Control (2020), and have been utilized by many organizations (including the Federal Reserve, Johnson & Johnson, Hartford Healthcare) worldwide for pandemic planning.

We then build models to calculate the various dimensions of costs given the realized pandemic. In particular, humanitarian costs include the costs due to loss of life, hospitalization, ICU, and ventilation incurred in the pandemic but also includes the psychological toll on the general population due to anxiety, PTSD, and depression. The economic costs take into account both the losses due to reduced output but also the increased economic burden due to unemployment. We construct a model for each individual cost item based on the latest studies.

To demonstrate the wide applicability of the THEMIS framework, we implement THEMIS for Germany, United States, Singapore, Spain and Brazil using the latest economic and health data (see Appendix B for sources).

Our experimental results yield three main implications. First, our results show that different governments face vastly different cost tradeoff curves in the pandemic. In particular, developing countries tend to suffer more under restrictive NPIs compared to developed countries, due to differences in economy composition, demographics, compliance, population density, healthcare infrastructure, among others. This suggests that developed countries potentially should take more burden of implementing restrictive policies in order to control the pandemic effectively, given the ineffectiveness of developing countries to do so. Second, we see that across countries mild pandemic restrictions are ineffective at the start of the pandemic and often produce the worst outcomes in terms of total cost. Contrary to many situations in which a moderate policy is preferred, we show that for a pandemic, moderate policies generate significant societal costs while are usually completely ineffective in stopping the spread of the pandemic. This provides further evidence that the lockdown policies widely adopted at the start of the pandemic were indeed beneficial, even when taking into account the costs of lockdown. Our final result highlights the outsized impact of when interventions start – we show that the total cost of the pandemic could decrease by 90% just by implementing policies one week earlier. This highlights the necessity for fast response in a pandemic, and also again reinforces how the exponential nature of the pandemic makes good policy-making difficult.

In summary, this paper makes two contributions. From a modeling standpoint, it formulates an original, modular system dynamics model for conducting cost-benefit analyses of COVID-19 NPIs. The model is capable to provide data-driven insights on how the pandemic could have evolved under different intervention scenarios, which is critical for policymakers to prepare for the next pandemic. The modular nature means that THEMIS could be easily applied to other regions. To facilitate this process, we open-source the THEMIS codebase and it can be found at https://github.com/COVIDAnalytics/THEMIS along with detailed instructions for reproducing and extending the results.

From a practical standpoint, this paper also consolidates the insights of applying the THEMIS model to a large range of countries during the first-wave of the pandemic. In particular, it demonstrates the highly complex nature of decision making in a pandemic to balance different dimensions of social cost. Further, it highlights how swift and strong action in the pandemic is the key to minimizing the total impact, and how different governments can optimize their strategies in the future.

## 2 THEMIS Framework Formulation

The schematic for the THEMIS Framework is shown in Figure 1. THEMIS starts with the input of a policy ***P*** = (***I, T***), which we formally define as a combination of *k* NPIs ***I*** = (*i*_0_, …, *i*_*k*−1_) and (*k* + 1) implementation times ***T*** = (*t*_0_, *t*_1_, …, *t*_*k*_) where *t*_0_ *< t*_1_ *<* … *< t*_*k*_. The policy is implemented such that each NPI *i*_*l*_, *l* ∈ {0, …, *k* − 1} is effective between [*t*_*l*_, *t*_*l*+1_], and is selected from a set of possible NPIs ℐ. For example, the policy ***P*** with:

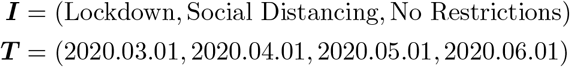

represents a policy from March 1st, 2020 to June 1st, 2020 with the stages as outlined in Table 1. We note that this structure is general and all policies implemented in the COVID-19 pandemic can be written in such structure, given a sufficiently large set of 𝒩. For simplicity only, we would assume throughout this paper that *t*_*l*_ for *l* ∈ {0, …, *k*} has units of days.

**Table 1:** Example Policy

| Policy Start Date | Policy End Date | Policy |
| --- | --- | --- |
| March 1st, 2020 | April 1st, 2020 | Lockdown |
| April 1st, 2020 | May 1st, 2020 | Social Distancing |
| May 1st, 2020 | June 1st, 2020 | No Restrictions |

**Fig. 1:**
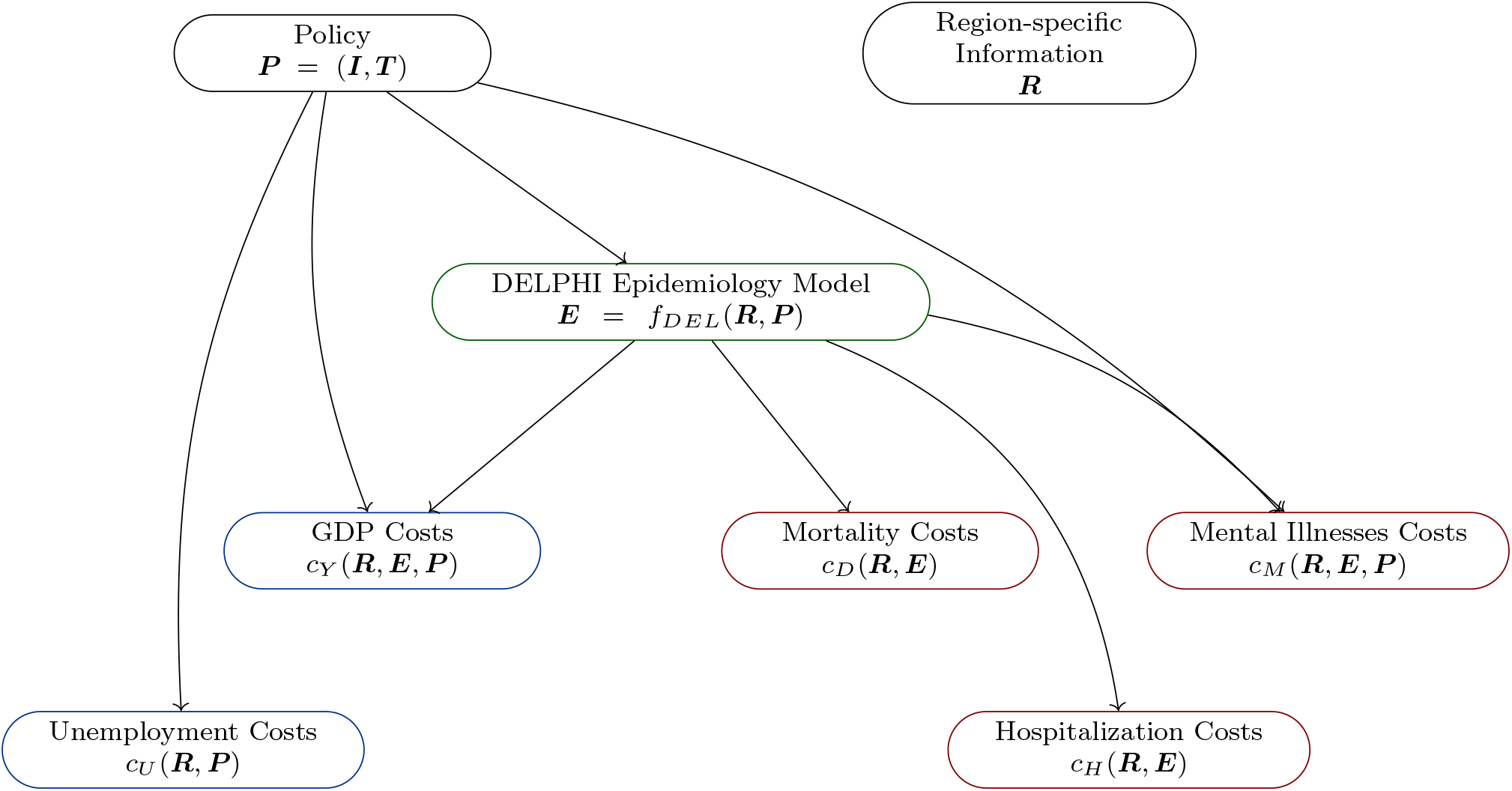
THEMIS Schematic. Here we ignore the arrows from ***R*** to every module for simplicity.

Now the goal of THEMIS is to calculate, for a specific region with region-specific data ***R***, the cost of such policy ***P*** within the implementation period *t* ∈ [*t*_0_, *t*_*k*_]. As illustrated in Figure 1, the first step is to utilize the DELPHI epidemiological model to simulate the spread of the epidemic ***E*** under such policy for this region ***E*** = *f*_*DEL*_(***R, P***). Then, we calculate the cost of the epidemic using both ***P*** and ***E***. In particular, we separated the cost into two categories, with five dimensions in total: economic cost (unemployment (*c*_*U*_), GDP (*c*_*Y*_)), and humanitarian cost (deaths (*c*_*D*_), hospitalizations (*c*_*H*_), and mental illnesses (*c*_*M*_)).

The following subsections describe the detail construction of each model (*f*_*DEL*_,*c*_*U*_, *c*_*Y*_, *c*_*D*_,*c*_*H*_, *c*_*M*_).

### 2.1 The DELPHI model: Forecasting the dynamics of the COVID-19 pandemic

DELPHI is a policy-driven compartmental epidemiological model that extends the widely used SEIR model to account for effects specific to the COVID-19 pandemic. The model is governed deterministically by a system of ordinary differential equations (ODEs) involving 11 states: susceptible (*S*), exposed (*E*), infectious (*I*), undetected cases who will recover (*U*^*R*^) or die (*U*^*D*^), detected hospitalized cases who will recover (*H*^*R*^) or die (*H*^*D*^), detected quarantined cases who will recover (*Q*^*R*^) or die (*Q*^*D*^), recovered (*R*) and deceased (*D*). Within the hospitalized states (*H*), there are also helper states to govern patients that are in the ICU (*IC*) and patients who are ventilated (*V*). Since its conception in late March 2020, it has been successfully applied to more than 210 countries and regions worldwide with high accuracy, and is utilized by organizations including the Hartford Hospital system for pandemic planning. (Li et al. 2020)

DELPHI differs from other COVID-19 forecasting models (see, e.g. Kissler et al. 2020) by capturing three key elements of the pandemic:

— **Under-detection**: Many cases remain undetected due to limited testing, asymptomatic carriers, and detection errors. Ignoring them would underestimate the scale of the pandemic. The DELPHI model captures them through the *U*^*R*^ and *U*^*D*^ states.
— **Governmental and societal response**: Social distancing policies limit the spread of the virus. Ignoring them would overestimate the scale of the pandemic. However, if restrictions are lifted prematurely, a resurgence may occur. We define a governmental and societal response function *γ*(*t*), which modulates the infection rate and is parameterized as follows:

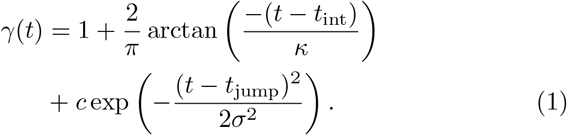

This parameterization encompasses four phases (Figure 2). In Phase I, most activities continue normally as people adjust their behaviors. This is followed by a sharp decline in the infection rate during Phase II as the policies get implemented. The parameters *t*_int_ and *κ* can be interpreted as the start time and the strength of this response. In Phase III, the decline in the infection rate reaches saturation. The epidemic then experiences a resurgence of magnitude *c* in Phase IV, due to relaxations in governmental restrictions and in social behaviors. This is counteracted at time *t*_jump_, when restrictions are reimplemented, with *σ* controlling the duration of this second wave.

**Fig. 2:**
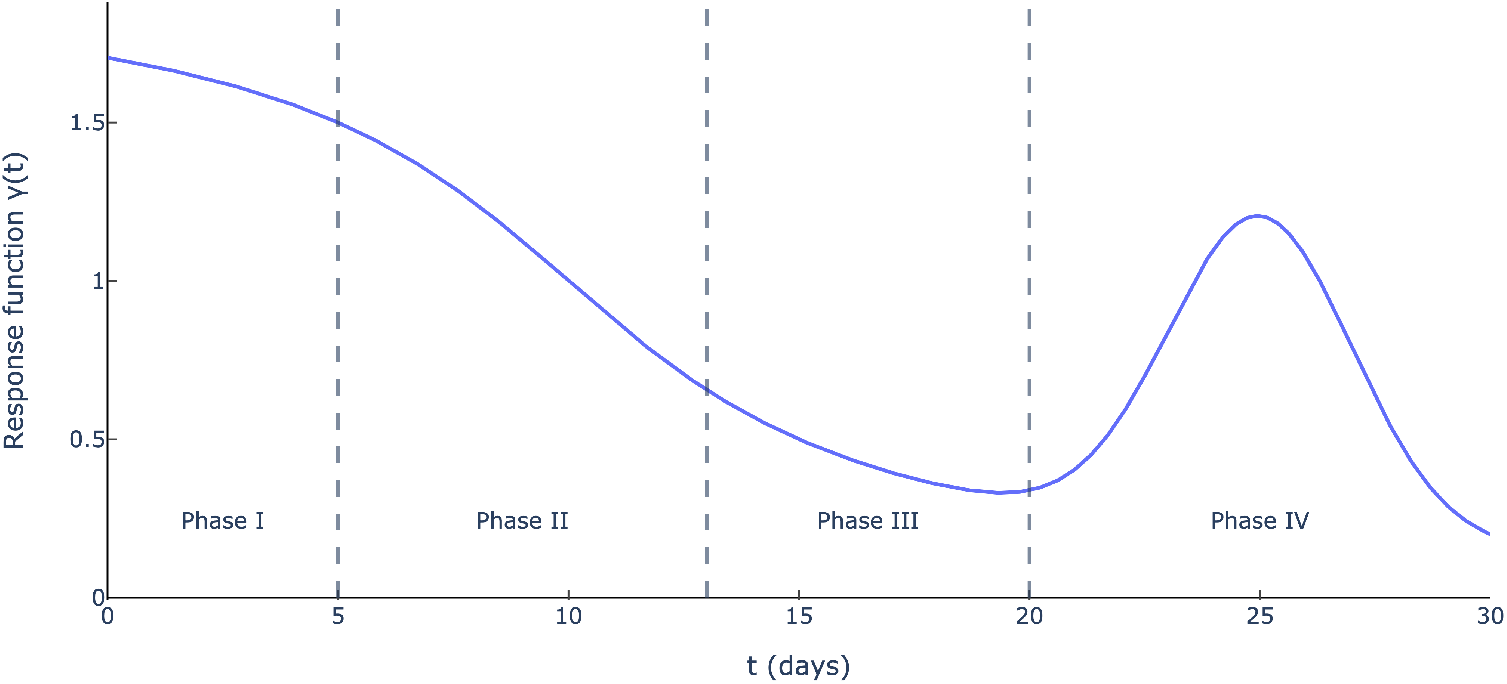
Governmental and societal response function *γ*(*t*) (*κ* = 5, *t*_int_ = 10, *c* = 1, *t*_jump_ = 25 and *σ* = 2).

— **Declining mortality rates**: The mortality rate of COVID-19 has been declining through the pandemic, due to a better detection of mild cases, enhanced care for COVID-19 patients, and other factors. We model the mortality rate as a monotonically decreasing function of time:

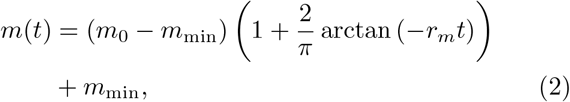

where *m*_0_ is the initial mortality rate, *m*_min_ is the minimum mortality rate and *r*_*m*_ is a decay rate.

Ultimately, DELPHI involves 16 parameters that define the transition rates between the 11 states. We calibrate 7 of them from a database on clinical outcomes (Bertsimas et al. 2020b). Using non-linear optimization, we estimate the other 9 parameters from historical data on the number of cases and deaths in each region. We provide the full mathematical formulation of the DELPHI model and its fitting procedure in Appendix A.

We utilize the DELPHI model to provide THEMIS with a highly accurate epidemiological model that responds to changes in policy. The key feature of DELPHI that is specifically relevant to THEMIS is that the rate of people being exposed to the virus and leaving the susceptible state (*S*) is policy-driven, and governed by the following differential equation:

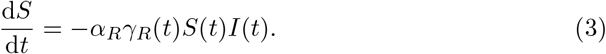

Here *α*_*R*_ can be interpreted as the natural infection rate of the epidemic in the region, while *γ*_*R*_(*t*) is the time-varying reduction of such infection rate due to the policies implemented in the region. *α*_*R*_ is extracted directly from the appropriate fitted parameters for DELPHI. For the actual policy implemented in the region, We can also estimate *γ*_*R*_(*t*) directly by the DELPHI model.

To create *γ*_*R*_(*t*) for a hypothetical policy ***P***, we first estimate a global correction factor *γ*_*i*_ for every potential NPI *i* ∈ ℐ using historical data (we assume that every *i* ∈ ℐ has been historically implemented somewhere). Specifically, *γ*_*i*_ is calculated as the average reduction of the natural infection rate *α*_*R*_ as observed all days and regions that *i* was implemented. We normalize these values so that *γ*_*i*_ = 1 for *i* = “No Measure”. Note that we are not using these correction values in our model directly but we use these to calculate the region specific *gamma*_*R*_ as explained below, which means implementing no NPI can also have a correction factor not equal to 1 for a region.

Then, for any region *R*, we calculate 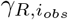 for every potential NPI *i*_*obs*_ ∈ ℐ that was implemented in the region by taking the average observed *γ*_*R*_(*t*) over all days that *i*_*obs*_ was implemented in region R. For a NPI *i*_*uobs*_ ∈ ℐ that were not historically implemented in region *R*, we perform a linear regression between the global and region-specific reduction factors 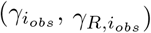, and utilize the imputed value as the region-specific reduction factor 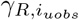, as shown in Figure 3.

**Fig. 3:**
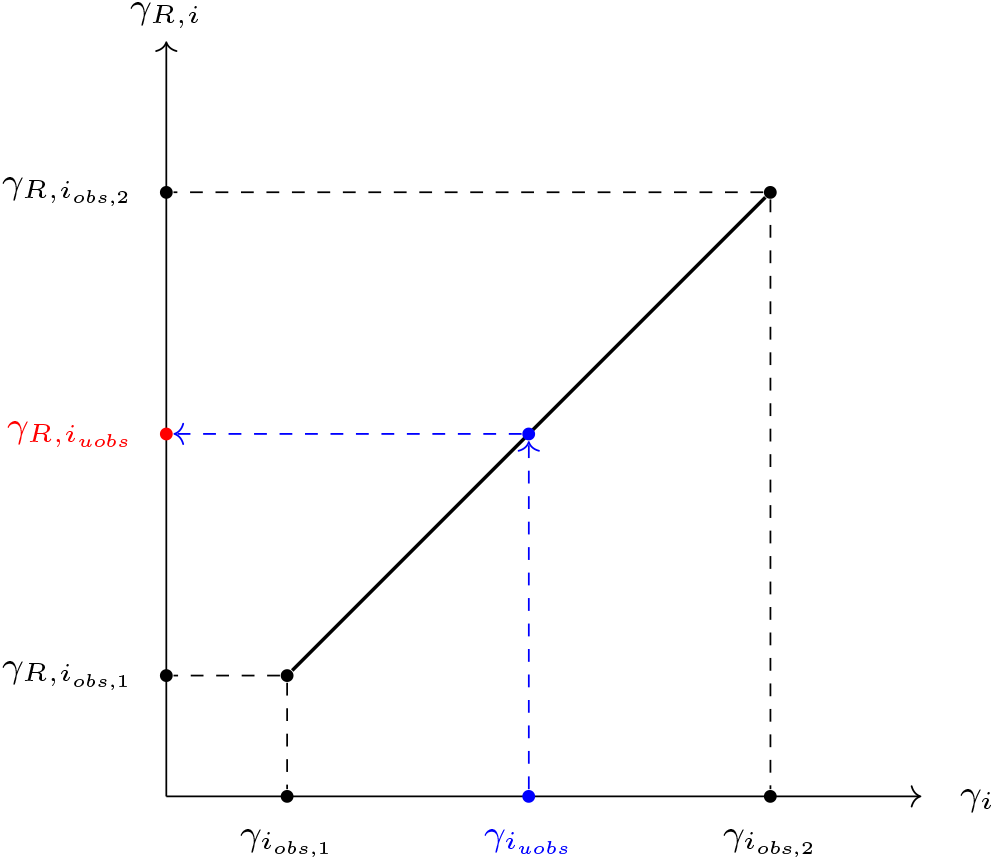
A linear interpolation procedure to impute the impact of policies on the infection rate 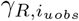 that were not implemented in region *R*, using the global estimated impact *γ*_*i*_.

Then, for a hypothetical policy ***P*** = (***I, T***), we define *γ*_*R*_(*t*) as a piecewise constant function of the region-specific reduction factors:

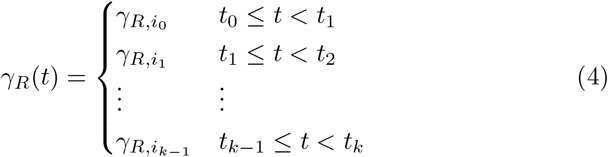

Intuitively, we are assuming that each NPI has a region-specific constant reduction on the infection rate during its duration of application. Thus, by changing the policy ***P***, the evolution of the pandemic ***E*** would be different.

### 2.2 Humanitarian Cost Models

The first major category of cost during the pandemic is the humanitarian cost. This includes not just the physical health component of hospitalization and deaths in the COVID-19 pandemic, but also the impact on mental health. The NPIs induce isolation and confinement, leading to increased mental illnesses, while healthcare workers on the COVID-19 front-line experience PTSD (Liu et al. 2020a, Carmassi et al. 2020). We develop models for these components of humanitarian cost below.

#### Cost of COVID-19 Deaths

We calculate the cost of COVID-19 deaths by considering the total quality-adjusted life years (QALYs) lost due to the premature mortality. This allows us to adjust for the skew in COVID-19 mortality that is predominantly concentrated in the elder age group. Specifically, we assume that each individual of age *u* who died from COVID-19 would have otherwise on average lived for *l*(*u*) years, where *l*(*u*) is the expected remaining life expectancy at age *u* for the population in the region under consideration (we omit the dependence on the region in the notation here for simplicity). Then, we denote the Probability Density Function (PDF) of age in COVID-19 deaths in that region as *f*(*u*). Using *f*(*u*) and *l*(*u*), we can calculate the expected QALY lost for each COVID-19 death as:

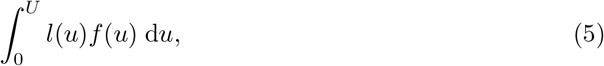

where *U* is some finite maximum age. Then, we calculate the total cost of QALYs lost using the total number of deaths in the epidemic *E*_*D*_ and the unit QALY cost *c*_*QALY*_ :

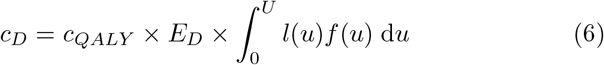

#### Cost of COVID-19 Hospitalizations

Our calculation of the cost of COVID-19 hospitalizations separates into three categories: general hospitalizations, Intensive Care Unit (ICU) hospitalizations, and ventilated hospitalizations. This separation is natural due to the large differences in both the length and cost of treatment across the three categories, of which the DELPHI epidemiological model takes into account. Therefore, we can extract the total number of hospitalization-days in each category from the DELPHI simulated pandemic ***E*** as *E*_*H*_, *E*_*I*_, *E*_*V*_ respectively. Then, denoting the daily treatment cost in each category as 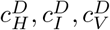, the cost of COVID-19 hospitalizations can be written as:

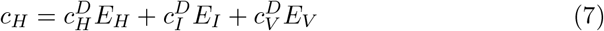

#### Cost of Mental Illnesses

For mental illnesses, we focus on the effects of the pandemic and policies on post-traumatic stress disorder (PTSD) and depression, highlighted by multiple studies as the leading mental health issues arising in the pandemic (e.g. Pfefferbaum and North 2020, Usher et al. 2020, Zhong et al. 2021). Specifically, there has been a sharp increase in PTSD among healthcare workers and hospitalized COVID-19 patients, while the general population experiences a significant increase in rate of clinical depression due to isolation caused by containment policies (Liu et al. 2020a, Carmassi et al. 2020). We would consider both these effects in our calculations. Specifically, we write *N* for the total population in the region, and *N*_*H*_ as the total number of health workers exposed to COVID-19.

To calculate the cost of depression, we make a few practical assumptions. First, we assume that the increase in clinical depression is only significant if the current NPI implemented is in the set of ”severe NPIs”, ℐ_*S*_ ⊂ ℐ, such as lockdown. Then, denote the increase in prevalence of clinical depression under severe NPIs in the general population as Δ*r*_*Dep*_ and the total general population as *N*. Then, if we write the daily cost of depression as *c*_*Dep*_, then the total cost of depression can be written as:

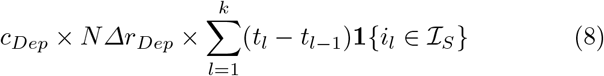

Compared to depression induced by the containment policies, PTSD due to either COVID-19 hospitalization or working closely with severe COVID-19 patients can be more persistent (Carmassi et al. 2020). We assume that PTSD due to effects of COVID-19 continue for *T*_*PTSD*_ days. We denote the increase in prevalence of PTSD among the susceptible population as Δ*r*_*PTSD*_. Then the total cost of PTSD can be written as:

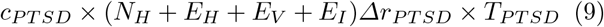

Therefore, the total cost of mental illnesses is:

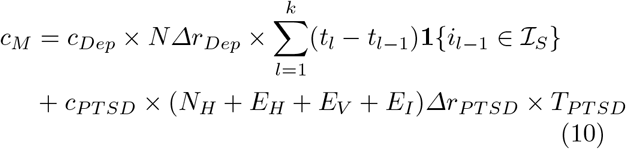

### 2.3 Economic Cost Models

Beyond the humanitarian impact, the COVID-19 pandemic created an outsized shock to the economy. The effect of the pandemic, and the NPIs aimed to slow the spread created large economic consequences that goes beyond the direct GDP impact. In particular, the reduced output triggered massive unemployment, which carries significant costs on its own. Therefore, in this category, we would consider both the direct costs of the pandemic on output, and the indirect costs of unemployment.

#### GDP Costs

The COVID-19 pandemic affects the GDP in at least two important ways. The first is the direct output loss due to sick workers for their duration of illness. Note that the long-term output loss due to deaths of individuals have already been considered in the costs of COVID-19 deaths, and thus we would only focus on the short-term output loss due to recovered COVID-19 patients. Denote the average length (in days) of COVID-19 sickness in the region as *T*_*S*_, and the number of yearly working days as *T*_*W*_. Then we assume that the total number of people who were sick, *E*_*S*_, is representative of the entire workforce, which has size *N*_*W*_. The total output loss due to sick workers is thus the appropriate portion of the entire yearly GDP, GDP_*Y*_:

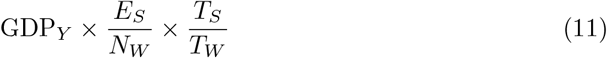

The second important mechanism in which the COVID-19 pandemic affects output is through the government policy ***P***. Concretely, we need to estimate a function Δ*Y*_*R*_(*i*) which outputs the impact on the annual GDP for region *R* for any NPI *i* ∈ ℐ. We do so by breaking down GDP of region *R* into private consumption (*C*), investment (*I*), government expenditures (*G*), and net exports (*NX*):

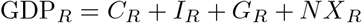

Then, we first establish the counterfactuals for each of the 4 constituents of annual GDP if the pandemic did not happen: *C*_*R*,0_, *I*_*R*,0_, *G*_*R*,0_, *NX*_*R*,0_. Then for a NPI *i*_*obs*_ ∈ ℐ that was implemented in region *R*, we calculate Δ*Y*_*R*_(*i*) as:

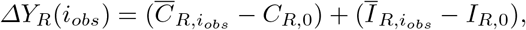

where 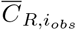 is the average annual private consumption over all periods during the pandemic when NPI *i*_*obs*_ was implemented, and similar for 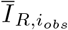. Note that we are ignoring the GDP change due to changes in government expenditure and net exports. We ignore changes in *G* to avoid the confounding effect due to government stimulus policies in the pandemic (which would not have likely been enacted if the pandemic had not occurred). We further ignore any fluctuations in *NX* as net exports are primarily affected by international policies, and therefore should not be counted towards the effect of *i* as applied within the region.

For NPI *i*_*uobs*_ ∈ ℐ that were not historically implemented in region *R*, we estimate Δ*Y*_*R*_(*i*_*uobs*_) by linear interpolation between 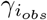 and Δ*Y*_*R*_(*i*_*obs*_) to impute Δ*Y*_*R*_(*i*_*uobs*_), similar to the procedure illustrated on Figure 3.

With Δ*Y*_*R*_(*i*), we can calculate the total output loss due to government policies as:

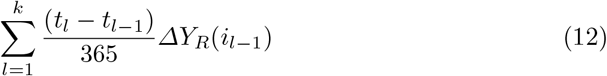

Therefore, the total GDP costs can be calculated as:

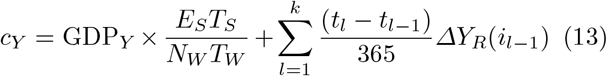

#### Unemployment Costs

Due to government-issued NPIs, many industries were forced to ground to a halt or severely reduce its capacity to produce, driving massive unemployment. In this model, we would calculate the *indirect* costs of unemployment beyond the direct loss of GDP as included in the model for GDP costs. Specifically, this refers to the costs of reduced well-being (physical and psychological) due to loss of work. We denote the yearly indirect cost of unemployment as 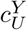.

Then similar to the GDP costs, we would estimate the function Δ*U*_*R*_(*i*) that measures the impact on unemployment rate for any NPI *i* ∈ ℐ. First, we establish a pre-pandemic unemployment rate *U*_*R*,0_ by averaging the seasonally-adjusted unemployment rate in region *R* 6 months prior to the pandemic. Then, for an observed NPI *i*_*obs*_ ∈ ℐ, we calculate Δ*U*_*R*_(*i*_*obs*_) as the average gain in unemployment rate over *U*_0_ across all months where *i*_*obs*_ was implemented. For an unobserved NPI *i*_*uobs*_ ∈ ℐ in region *R*, we again utilize linear interpolation with *γ*_*i*_ to impute Δ*U*_*R*_(*i*_*uobs*_), similar to the procedure illustrated on Figure 3. Then the total cost of unemployment can be calculated as:

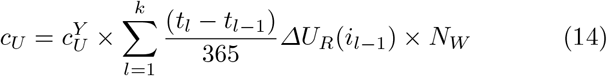

## 3 Analyzing the Governmental Response in the First Wave of the COVID-19 Pandemic

Using the THEMIS model, we now proceed to analyze the governmental response during the COVID-19 pandemic. As a demonstration, we focus on the first wave as it spread around the world (*t*_0_ ≥ 2020.03.15, *t*_*k*_ ≤ 2020.06.15). We aim to understand the costs of the pandemic induced by the actual policy and compare to alternative strategies to understand how different countries could have better responded to the pandemic.

### 3.1 Data and Experimental Setup

During the first wave of the pandemic, there was still much unknown about COVID-19. Although the Wuhan outbreak proved COVID-19’s capability to transmit between humans, key characteristics such as symptoms, mode of transmission, and mortality, were still being heavily debated in the scientific community. Therefore, most countries around the world focused on NPIs that primarily affected population mobility, including restrictions on travel, work, school, or stay-at-home/lockdown orders.

With such context, in the THEMIS model simulation, we would limit the set of potential NPIs ℐ to only mobility restrictions that were actually implemented around the world during the very early stages of the first wave (2020.03-2020.04). This ensures that our comparison is realistic and we do not propose alternative strategies that rely on information unavailable during the early stages of the pandemic.

We collected the mobility restriction NPIs deployed by 167 countries and regions around the world during March and April of 2020, and categorized the NPIs based on whether they restrict mass gatherings, schools, travel and work activities. We group travel restrictions and work restrictions together due to their tendency to be implemented simultaneously. The set ℐ includes six strategies, ranked from least to most severe: (1) *No measure*; (2) *Restrict mass gatherings only*; (3) *Restrict mass gatherings, travel and work*; (4) *Restrict mass gatherings and school*; (5) *Restrict mass gatherings, schools, travel and work*; and (6) *Stay-at-Home /Lockdown*. We consider the last two NPIs as “severe NPIs” (*I*_*s*_), incurring costs due to mental illnesses. We specify that each NPI would last for one month, and thus throughout the period of consideration [2020.03.15, 2020.06.15] each policy consists of three different NPIs. We consider all 6^3^ combinations of NPIs as valid policies ***P***. For simplicity, we would refer to policies by the numerical numbering of the NPIs below - a 6-6-6 policy thus represents a policy that enforced lockdowns for 3 months.

To demonstrate the wide applicability of THEMIS, we apply it to a diverse selection of countries around the world: Germany, United States, Singapore, Spain and Brazil. To ensure the greatest accuracy in modeling the alternative strategies, we take the region-specific epidemiological parameters from the DELPHI model trained at the end of the first wave (2020.07.01), and the region-specific reduction coefficients *γ*_*R,i*_ are estimated using the process detailed in Section 2.1. For the region-specific cost parameters, we take the most recent available data, and apply an appropriate inflation correction if necessary. The full table is included in Appendix B.

### 3.2 Results and Discussion

In Figure 4, we present the results of the simulation of all valid policies ***P*** for the countries specified above. We graph the resultant cost of such policy along both the humanitarian dimension (including mortality, hospitalization and mental illnesses’ costs), and the economic dimension (including GDP and unemployment). The cost of the actual implemented policy is denoted in blue. We immediately observe that the different regions have drastically dissimilar tradeoff curves, which highlight that a policy suitable for a certain area may very well be suboptimal for another. This is a reflection of the significant differences in cultural and governmental structure and capabilities across the different regions analyzed here.

**Fig. 4:**
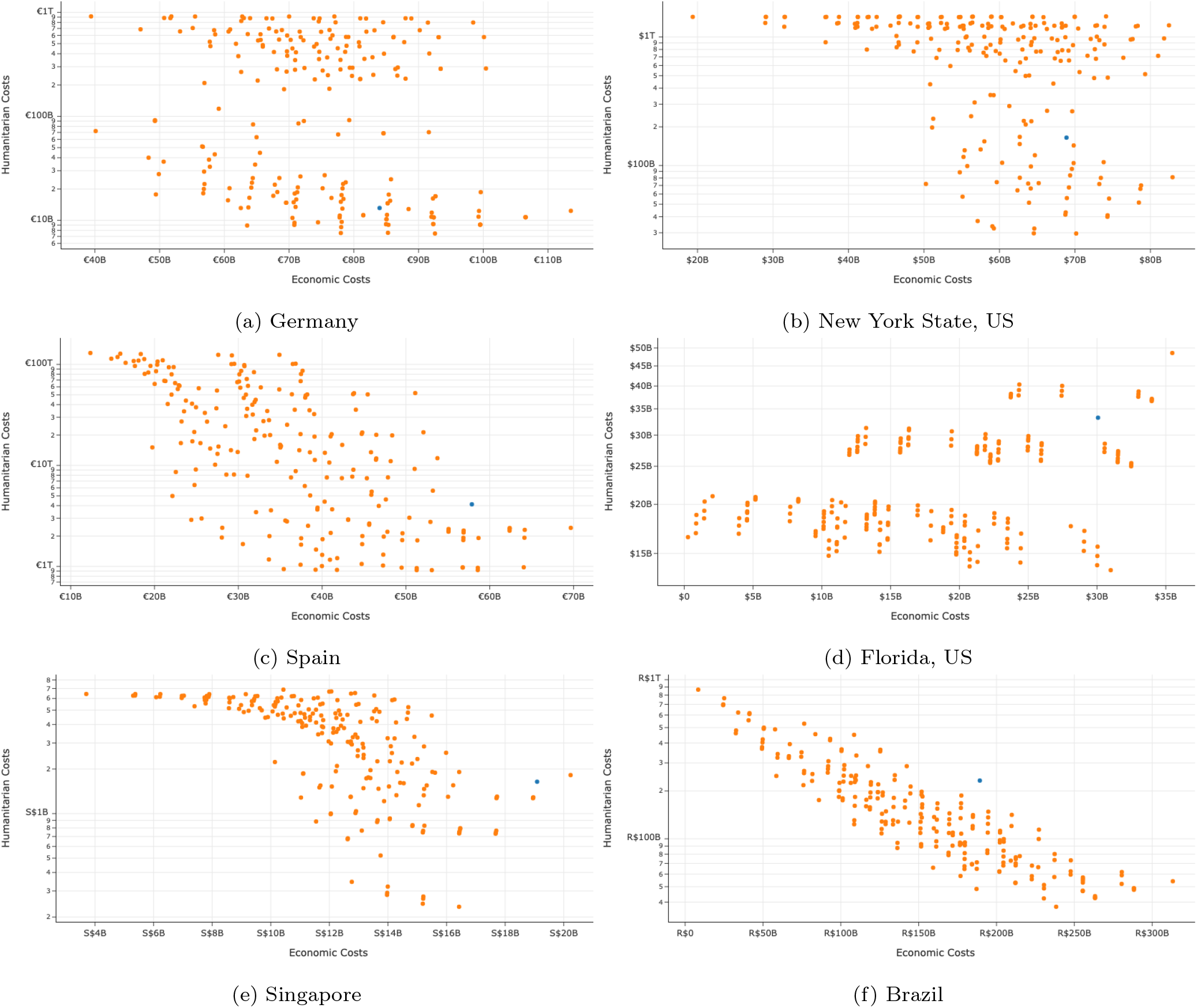
Tradeoff of Humanitarian and Economic Costs for various governmental policies ***P*** starting on March 15th and continuing for 3 months. Blue indicates the cost of the actual policy implemented over such period.

However, despite the immense differences, there are still some important general trends that we can draw from these graphs. First, we see that the incurred humanitarian cost varies logarithmically while the economic cost roughly stays on the same order of magnitude. This is a reflection of the exponential nature of a pandemic - NPIs insufficient to control the spread of COVID-19 carry exponentially more humanitarian cost than those that do. We empirically observe that policies with the highest humanitarian cost most often are policies that assume a very relaxed measure during the start of a pandemic (e.g. Restrict Mass Gatherings only), regardless of the policies later in the pandemic. This demonstrates the outsized importance of policy timing in regards to controlling the pandemic. In contrast, most policies seem to incur between 10 − 30% of the GDP of that region within the 3-month period.

Therefore, the ”efficient frontier” of the policies in terms of economic and humanitarian costs in instituting NPIs is very flat: An exponential decrease in humanitarian cost only requires a small increase in economic costs. For example, for Germany, compared with the 1-1-1 policy, the 6-6-6 policy (that institutes 3 months of lockdown) reduces humanitarian costs by over 1 Trillion Euros with only an increase in economic costs of less than 70 Billion. We can also compare the efficient frontier with the actual policy implemented in the regions, demonstrated by the blue dots in Figure 4, to understand how different policymakers valued the tradeoffs differently. In Germany, we see that the actual policy implemented is very close to the minimum humanitarian cost achievable, while the economic costs are higher. This suggests that the German government implemented a policy that highly valued a reduction in humanitarian cost. We also note that the actual policy is relatively close to the efficient frontier, suggesting that the German government had implemented a very successful policy in combating the first wave of the COVID-19 pandemic. In contrast, the actual policy for New York is quite far away from the efficient frontier, incurring a significant economic cost yet still registering a high humanitarian cost. The optimal policy for New York could have saved 120 billion dollars in estimated humanitarian cost while *also* saving 4 billion dollars in estimated economic cost.

However, the presence of an efficient frontier that is heavily biased towards reducing humanitarian costs does not mean that a more severe NPI is always better. In fact, across all countries tested, the most severe policy, 6-6-6, is never the optimal policy in terms of achieving minimum costs in the sum of the two dimensions.

The Figure 5 shows 20 minimum overall cost policies for all regions and the breakdown of all the costs. It shows how the share of all cost components for the optimal policies of different regions are so different. For example, for Germany, the optimal policies have very low *Loss of Life Costs* (about €10 billion) but higher share of *Economic Costs* (about €60 billion). On the other hand, for Spain, even with a more severe policy of 5-5-3 the *Loss of Life Costs* are the dominant costs at around €920 billion out of €960 billion, which justifies taking those more severe policy measures for Spain.

**Fig. 5:**
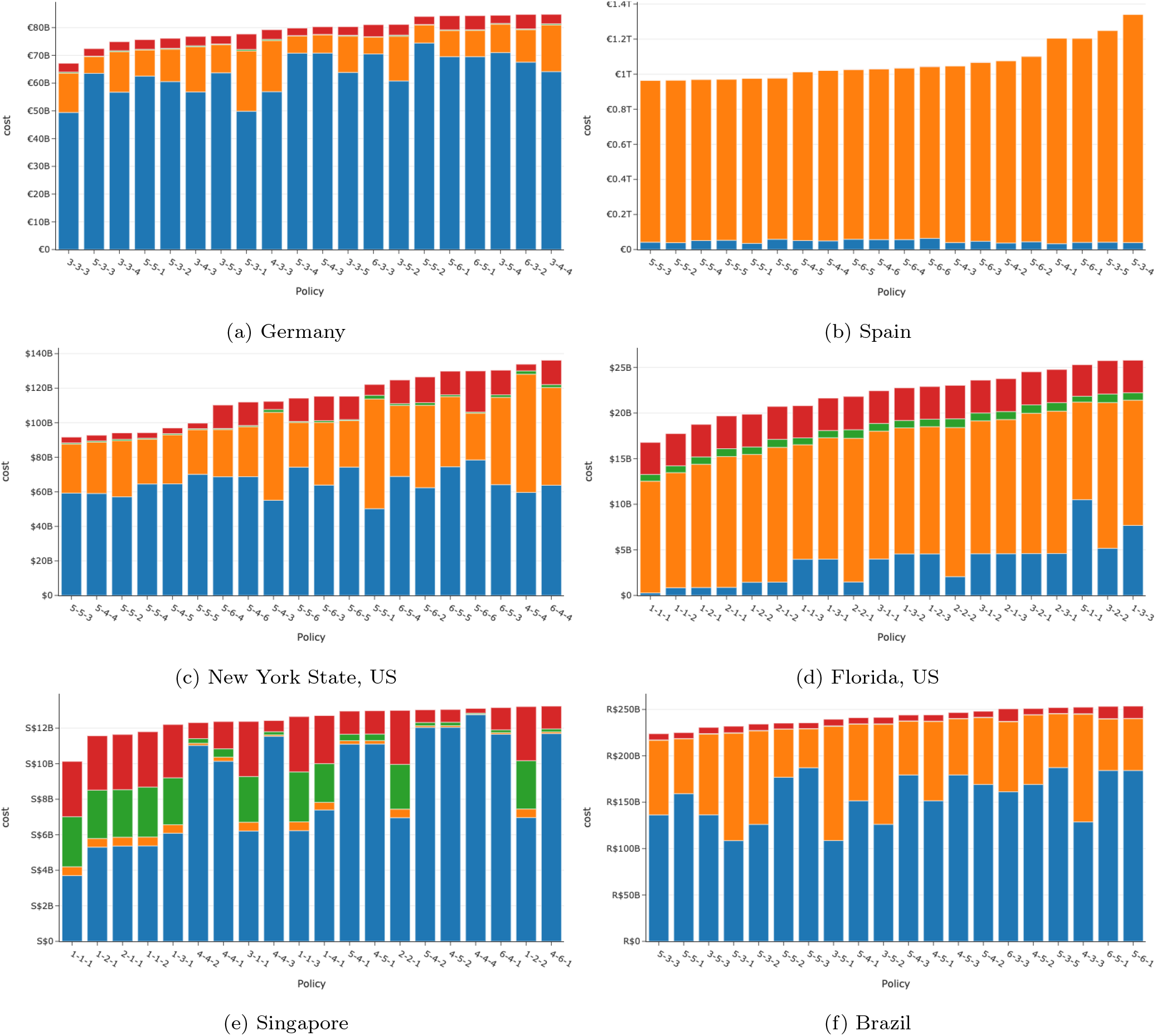
The 20 minimum total cost policies for different regions, starting on March 15th and continuing for 3 months. The red bar is mental health costs, the orange bar is loss of life costs, the green bar is hospitalization costs, while the blue bar is economic costs.

For New York State, Spain, and Brazil, the optimal policy is to have a strong initial response, followed by slow gradual reopening can avoid the vast majority of humanitarian costs while also protecting the economy. In contrast, in Florida and Singapore, the most optimal policy is to institute no measures for 3 months (1-1-1). This is because in both regions, the model believes that additional restrictions would bring little benefit. In Florida, the model estimates that the pandemic is already so widespread that additional restrictions, leading to large economic costs, would not save many additional lives. In Singapore, the model predicts that there is not enough local transmission to sustain an epidemic even if no restrictions were instituted. This again highlights the critical importance of timing for these government actions.

Furthermore, we observe that regions with high GDP per capita (US, Singapore and Germany) have different behavior with regions with lower GDP per capita (Spain and Brazil). For regions with high GDP per capita, we note in Figure 4 that generally there is a ”twin peak” structure - the policies with the highest economic costs tend to occur both at those with the least humanitarian costs, but also those with the highest humanitarian costs. The peak at the minimum humanitarian costs are a result from policies that continually apply the most severe NPIs, culminating in a large reduction in societal output. The peak at the maximum humanitarian costs are due to an uncontrolled pandemic reducing worker availability across the society, and thus GDP.

In contrast, for Spain and Brazil, high humanitarian costs does not mean high economic costs (due to reduced worker availability). This is because even if the government implements a relaxed COVID-19 policy, the effect of reduced output caused by COVID-19 infections is less significant in areas with lower GDP per capita. This indicates that developing countries might have more economic incentive to institute a more relaxed pandemic policy.

## 4 Limitations

Although the THEMIS model uncovered important actionable insights for policymakers, there are some innate limitations. First, due to the data-driven nature of estimating the effect of NPI, the THEMIS framework is unable to analyze any alternative NPI *i* that has not been historically implemented. This in turn restricts the potential set of alternative policies that we can consider.

Another important limitation of the THEMIS model relates to confounding in estimating the policy effect on the pandemic. Due to the observational nature of data, many societal variables change concurrently as NPIs are implemented, creating potential confounding effects. For example, voluntary behavior changes during the pandemic could inflate the perceived effect of NPIs on reducing infection rates. It is therefore important to attempt to isolate the effect of NPIs so that the counterfactual estimates are reliable. The THEMIS model, in its construction, attempts to minimize confounding whenever possible. For example, the DELPHI model contains many epidemiological parameters in attempt to isolate the policy-driven effect on the pandemic. The GDP cost model removes the change in GDP due to governmental expenditures and net exports as these are confounded by additional policies not within the scope of this paper. However, despite these measures, it is likely that residual confounding remains. Therefore, the conclusions drawn in this paper should be treated as a first step to attempt to create a holistic cost-benefit analysis of various policies. Further research is needed to develop a greater understanding of how the NPIs and other levers work together in affecting key dimensions of a society during a pandemic.

The modeling of various models within THEMIS also carries limitations of its own. The DELPHI model, while accurate, is still a simplification of the complex real-world dynamics of a pandemic. In particular, DELPHI carries the same limitations as other SEIR-based models in requiring a sufficiently large population and epidemic size for the large-scale population dynamics of compartmental models to be accurate. (Holmdahl and Buckee 2020) This means that the THEMIS model might be unsuited for application to granular regions or to regions where the epidemic is not yet significant. Furthermore, in many of the cost models, we have assumed the effects are linear and additive, when in reality non-linearities are often present. These assumptions were necessary to create a tractable model that could be supported by available data.

## 5 Conclusion

In this paper, we presented a system dynamics framework, THEMIS, that allows us to compare both the humanitarian and the economic effects of different NPIs on the society during a pandemic. THEMIS builds upon a state-of-the-art epidemiological model, DELPHI, and constructs data-driven cost models to analyze the impact of NPIs across mortality, hospitalizations, mental illnesses, GDP, and unemployment. We applied THEMIS to a wide variety of countries; the results demonstrate that early application of severe NPIs for a short period of time generally minimized total societal cost but the situation differs widely between countries. In particular, we note that developing countries face a relatively higher cost in implementing severe NPIs. The THEMIS framework is open-source and can be easily extended to other countries/regions around the world.

## Data Availability

All code and data are available online at https://github.com/COVIDAnalytics/THEMIS.

## A The DELPHI Model

The DELPHI model is a compartment epidemiological model that extends the classical SEIR model into 11 states under the following 8 groups:

— **Susceptible (*S*)**: People who have not been infected.
— **Exposed (*E*)**: People currently infected, but not contagious and within the incubation period.
— **Infected (*I*)**: People currently infected and contagious.
— **Undetected (*U***_***R***_**) & (*U***_***D***_): People infected and self-quarantined due to the effects of the disease, but not confirmed due to lack of testing. Some of these people recover (*U*_*R*_) and some die (*U*_*D*_).
— **Detected, Hospitalized (*DH***_***R***_**) & (*DH***_***D***_): People who are infected, confirmed, and hospitalized. Some of these people recover (*DH*_*R*_) and some die (*DH*_*D*_).
— **Detected, Quarantine (*DQ***_***R***_**) & (*DQ***_***D***_): People who are infected, confirmed, and home-quarantined rather than hospitalized. Some of these people recover (*DQ*_*R*_) and some die (*DQ*_*D*_).
— **Recovered (*R*)**: People who have recovered from the disease (and assumed to be immune).
— **Deceased (*D*)**: People who have died from the disease. In addition to main functional states, we introduce auxiliary states to calculate a few useful quantities: Total Hospitalized (TH), Total Detected deaths (DD) and Total Detected Cases (DT). The full mathematical formulation of the model is as followed:

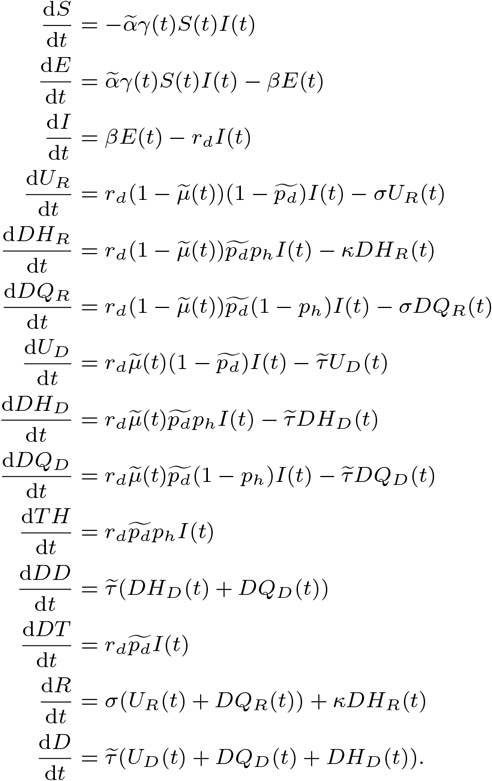

Figure 6 depicts a flow representation of the model, where each arrow represents how individuals can flow between different states. The underlying differential equations are governed by 16 parameters which are shown on the appropriate arrows in Figure 6 and defined below. To limit the amount of data needed to train this model, only the parameters denoted with a tilde are being fitted against historical data for each area (country/state/province); the others are largely biological parameters that are fixed using available clinical data from a meta-analysis of over 190 papers on COVID-19 available at time of model creation (Bertsimas et al. 2020a). A small selection of references for each parameter is given below.

**Fig. 6:**
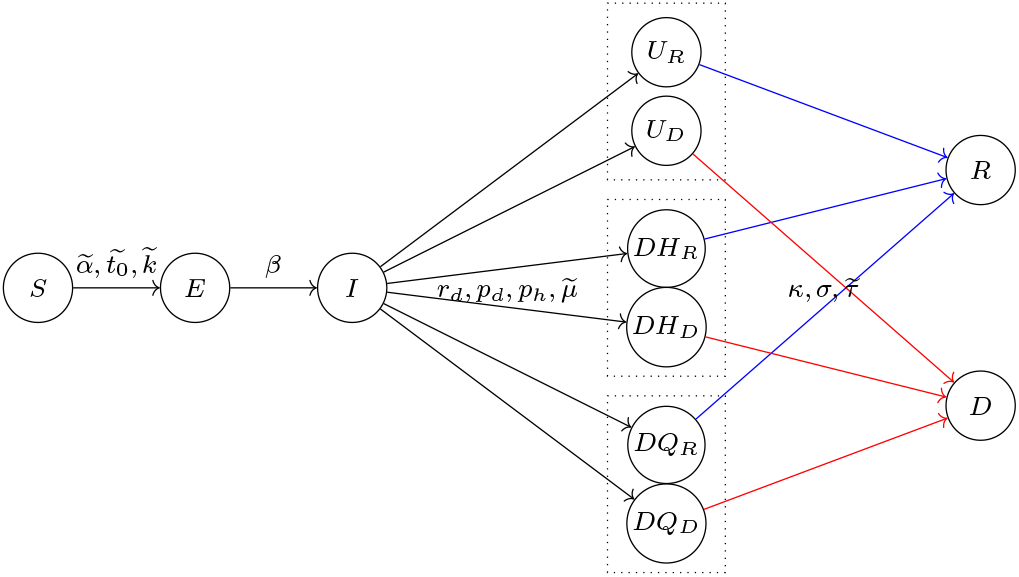
Flow Diagram of DELPHI.

— 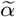 is the baseline infection rate.
— *γ*(*t*) measures the effect of government response and is defined as:

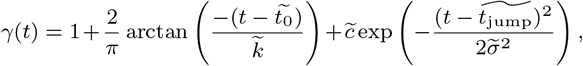

where the parameters 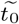 and 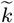 capture, respectively, the timing and the strength of the response. The exponential term intends to reflect a resurgence in infections due to relaxation of governmental policy and societal response, where 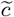 controls the magnitude of resurgence, 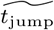 the time of the acme of the resurgence, and 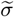 the duration of the resurgence phase. The effective infection rate in the model is 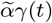, which is time dependent. The 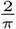 constant is so that the starting *γ*(*t*) with 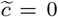 is normalized to the range of [0, 2] with *γ*(*t*) = 1 if *t* = *t*_0_.
— *r*_*d*_ is the rate of detection. This equals to 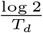, where *T*_*d*_ is the median time to detection (fixed to be 2 days), see Wang et al. (2020).
— *β* is the rate of infection leaving incubation phase. This equals to 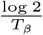, where *T*_*β*_ is the median time to leave incubation (fixed at 5 days), see Lauer et al. (2020).
— *σ* is the rate of recovery of non-hospitalized patients. This equals to 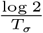, where *T*_*σ*_ is the median time to recovery of non-hospitalized patients (fixed at 10 days), see Hu et al. (2020), Kluytmans et al. (2020).
— *κ* is the rate of recovery under hospitalization. This equals to 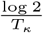, where *T*_*κ*_ is the median time to recovery under hospitalization (fixed at 15 days), see Liu et al. (2020b), Grein et al. (2020).
— 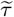 is the rate of death. This captures the speed at which a dying patient dies, and thus inversely proportional to how long a dying patient stays alive.
— 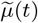 is the case fatality rate, defined as:

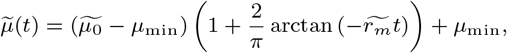
— Where 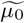 is the initial case fatality rate, *µ*_min_ is the minimum case fatality rate and 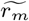 is the decay rate for mortality. This parametric curve describes the natural decay of case fatality rate as standard of care improves throughout the pandemic. Notice that this quantity measures the percentage of people who die from the disease in a particular region, and is independent from the rate of death. If *r*_*m*_ *<* 0, this function can also capture the effect of an increasing mortality rate.
— *p*_*d*_ is the percentage of infectious cases detected, which is fixed at 20% based on various reports trying to understand the extent of underdetection in countries with earlier outbreaks. Wang et al. (2020), Krantz and Rao (2020), Niehus et al. (2020)
— *p*_*h*_ is the (constant) percentage of detected cases hospitalized, which is set to 15%, see Arons et al. (2020), Xu et al. (2020).

We fit on 11 parameters from the list above 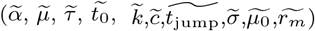. In addition, we introduce 2 additional parameters 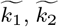 to account for the unknown initial population in the infected (*I*) and exposed (*E*) states. We thus fit 13 parameters per area.

The parameters are fitted by minimizing a weighted Mean Squared Error (MSE) metric with respect to the parameters. Let *DT* (*t*) and *DD*(*t*) denote the number of reported total detected cases and detected deaths, respectively, on day *t*. Then, the loss function for a training period of *T* days is defined as:

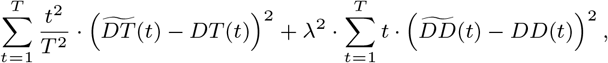

where 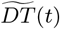 and 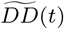 are respectively the total detected cases and deaths predicted by DELPHI. The factor 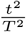 gives more prominence to more recent data, as recent errors are more likely to propagate into future errors. The lambda factor 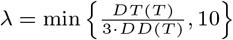 balances the fitting between detected cases and deaths; this re-scaling coefficient was obtained experimentally. We specifically exclude historical data starting before the area recorded more than 100 cases; this allows us to exclude sporadic outbreaks that are not epidemics. To optimize over the highly non-convex search space, we utilize both the local truncated newton algorithm (TNC) (Nocedal and Wright 2006) and the global optimization method of dual annealing (DA) (Xiang et al. 1997).TNC is utilized to produce forecasts on a daily basis while DA, being more computationally expensive, is performed on a weekly basis to shift and re-adjust the parameters more significantly if the underlying mechanics have changed (e.g. in the case of a new wave of cases). We use a bound of 20% deviation around the latest value for TNC, and a bound of 50% deviation for DA.

## B List of Region-specific Cost Parameters

**Table 2:**
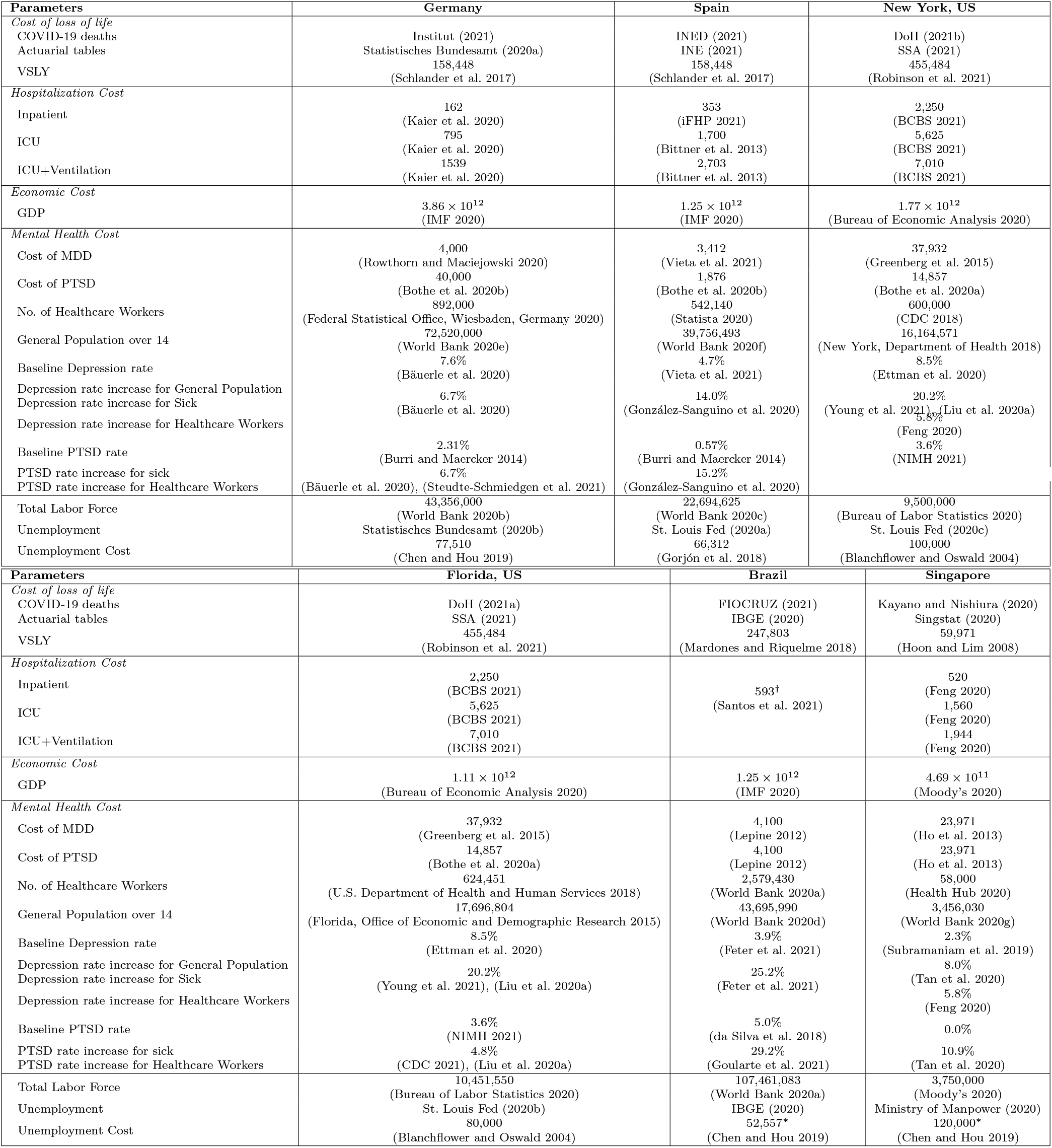
Values and sources for the THEMIS parameters used in Section 3. All monetary units are in local currency units unless otherwise specified. For sources with monetary information not from the simulation year (2020), we correct the monetary value using the Consumer Price Index. †We were only able to find the blended average hospitalization cost for COVID-19 across inpatient, ICU, and ventilated patients. ^∗^We extrapolate from the observation in the paper that unemployment cost is around 1.5 year of GDP per capita.

